# Evaluation of Peyton’s Four-Step Approach for Skill Acquisition in Undergraduate Medical Students: A Prospective Randomized Study

**DOI:** 10.1101/2023.07.27.23293240

**Authors:** Rohin Garg, Gaurav Sharma, Anil Chaudhary, Simmi Mehra, Poonam Sood Loomba, Vijendra Devisingh Chauhan

**Affiliations:** Department of Anatomy, All India Institute of Medical Sciences, Rajkot, Gujarat, India; Department of Physiology, All India Institute of Medical Sciences, Rajkot, Gujarat, India; Department of Microbiology, All India Institute of Medical Sciences, Rajkot, Gujarat, India; Microbiology, Maulana Azad Medical College and G B Pant Hospital, New Delhi, India; Swami Rama Himalayan University, Orthopaedic Surgery, Himalayan Institute of Medical Sciences, Jolly Grant, Dehradun, Uttarakhand, India

**Author notes:** **Corresponding Author:** Dr. Rohin Garg, Associate Professor, Department of Anatomy, All India Institute of Medical Sciences, Rajkot, Gujarat, India., Contact.

**Keywords:** Conventional teaching methods, MBBS, Peyton’s technique, Social Learning theory

## Abstract

**Background:** Skills laboratory training for procedural skills has gained a lot of significance in recent years. An instructional approach that is becoming increasingly prevalent in medical education is Peyton’s four-step approach which demonstrates benefit of peer learning under supervision of teachers.

**Aim:** The aim of this study was to determine the effect of application of Peyton’s four-step approach versus traditional learning on medical students’ performance.

**Objectives:** To compare the effectiveness of training effect of application of Peyton’s four-step approach versus traditional learning on medical students’ performance to measure blood pressure, and in performing hand washing procedures. As well as to assess and compare student self-satisfaction level and confidence level.

**Methodology:** The present prospective intervention study was carried out among 50 Medical students of Phase I MBBS by 4 faculty members with approval from the Institutional Ethics Committee. [Group A (intervention Group) and B (Control Group)], 25 students in each group were selected by using randomization. For the control group; the blood pressure measurement procedure and washing techniques were explained using the two-stage approach “see one, do one” and for the study group; each subgroup was trained to perform blood pressure measurement procedure and washing techniques using “Peyton’s four-step approach. A common scoring checklist for outcome evaluation was prepared to evaluate the student’s skills following the training session to avoid discrepancies in scoring among the faculties.

**Results:** It was found that in the control group 52% students were able to measure blood pressure and in study group 88% of students were able to do following skill sessions and in the control group 48% students were able to wash hands as per required protocol and in study group 96% of students were able to do following skill sessions. Regarding Students self-confidence and satisfaction levels, 24% were highly satisfied and confident in the control group as compared to 88% students among study group and a statistical difference was noted in confidence level in between two groups. Regarding the average and very good effectiveness of training, a statistical difference was found between control and study group.

**Conclusions:** Students who learnt through using Peyton’s four–step approach showed rapid learning, more confidence and exhibited higher competency in performing blood measurement procedure and hand washing techniques than those who learned through traditional method.

## BACKGROUND

Traditionally procedures are taught using a “see one - do one” approach which is known as Halsted’s teaching approach and was based on the surgeon Halsted (1904).(1,2) This technique refers that a teacher demonstrates and describes a procedure and afterwards the students are asked to practice the procedure. However, given the diversity of existing procedures today, others say that the teaching approach should be modified to “see many, learn from the result and do many” ^2^ Hence, Rodney Peyton’s four-step approach has been proven to be effective in skills lab training of technical skills.(3,4) The approach comprises four clearly defined instructional steps:(5)

- Step 1 – “Demonstrate”: The trainer demonstrates the skill at a normal pace and without additional comments.
- Step 2 – “Talk the trainee through”: The trainer demonstrates the respective skill while describing each procedural substep in detail.
- Step 3 – “Trainee talks trainer through”: The trainer performs the skill for a third time, based on the substeps described to him by the trainee.
- Step 4 – “Trainee does”: The trainee performs the skill on his/her own.

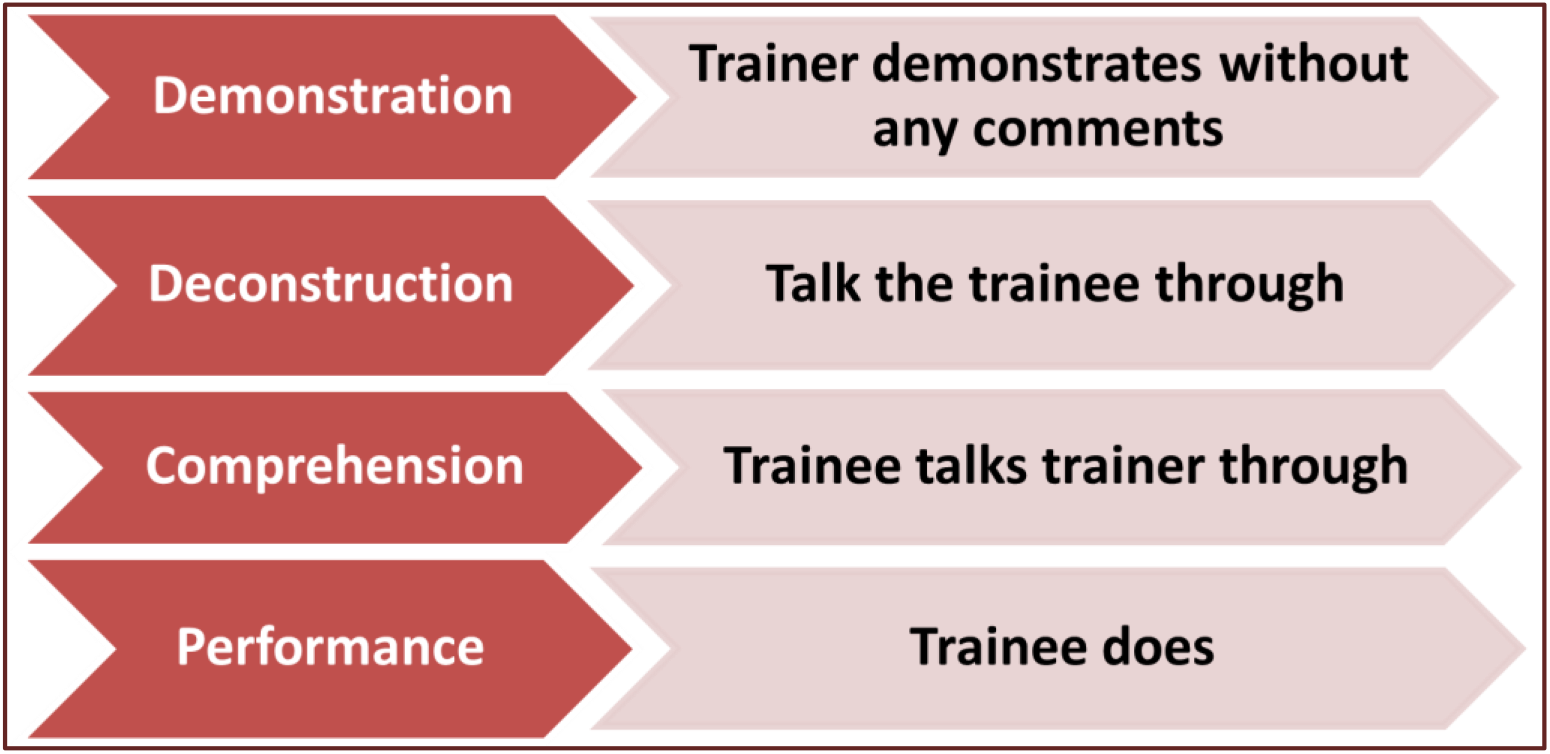

Payton’s 4 step approach has several advantages as the student has to think in steps 1 and 2 first before sharing the instruction with the teacher. This allows students to consolidate their ideas before actively expressing them. In addition, the intellectual process called self-explanation seems to simplify the amalgamation of innovative knowledge into existing knowledge.(6) Furthermore, as a benefit of peer learning, it seems necessary to appreciate friendship in clinical learning environments among undergraduate students.(7) This project aims to evaluate whether Peyton’s four-step approach is superior to conventional instruction for teaching skills to medical students and whether skills are maintained over time.

## AIM AND OBJECTIVES

### Aim

The aim of this study was to determine the effect of application of Peyton’s four-step approach versus traditional learning on medical students’ performance

### Objectives

i. To compare the effectiveness of training effect of application of Peyton’s four-step approach versus traditional learning on medical students’ performance to measure blood pressure.
ii. To compare the effectiveness of training effect of application of Peyton’s four-step approach versus traditional learning on medical students’ performance in performing hand washing procedures
iii. To assess and compare student self-satisfaction level in study group and control group.
iv. To assess and compare student confidence level in study group and control group.

## METHODOLOGY

### Study design

Prospective intervention study (Experimental educational approach)

### Study Setting

Department of Anatomy, AIIMS, Rajkot, Gujarat, India.

### Participants

Medical students of Phase I MBBS and 4 faculty members with approval from the Institutional Ethics Committee. [Group A (intervention Group) and B (Control Group)], equal students in each group were selected by using randomization.

### Sample size

50 medical students; [Group A (intervention Group) and B (Control Group)] of 25 students in each group.

### Sampling

By Complete enumeration, assuming that each participant has an equal chance of being assigned to any group. For randomization method, students were selected by lottery system.

### Description of Intervention

**For the control group**, the blood pressure measurement procedure was explained using the two-stage approach “see one, do one” in which the procedure was explained to the students by the clinical instructor only one time (demonstration) then they were allowed to do it independently (re-demonstration) under the instructor’s supervision.

**For the study group;** each subgroup was trained to perform blood pressure measurement procedure using “Peyton’s four-step approach” following four sequential steps; demonstration, deconstruction, comprehension, and performance as follow:

During the first step, the instructor demonstrated the procedure silently “at normal speed, and without commentary”**(Demonstration**).

Then, the instructor demonstrated the procedure while describing each step to the students (**Deconstruction)**.

During the third step, the instructor demonstrated the procedure following students’ instruction for each step (**Comprehension**)

Finally, students simultaneously demonstrated and described the procedure step by step (**Performance.)**

In this study, following skill sessions was conducted:

1. Blood pressure Measurement
2. Hand washing techniques

Students’ skill acquisition was measured for **both groups**; after performing the procedure of it using checklist.(8,9)

For Blood Pressure Measurement Technique, patient was prepared by asking patient if he/she has ingested caffeine or used nicotine within the past 60 minutes or exercised within the past 30 minutes. Also, it was noted if the patient is in pain or very emotionally upset. Patient was made to sit quietly for at least 5 minutes prior to measurement. The cuff was selected correctly and equipment was checked. The gauge needle or mercury column was checked if it was at zero. Patient/equipment were positioned correctly. Measurement was done on bare arm, if a sleeve was not rolled up, it was removed. Patient was seated in chair with back supported. Feet was placed flat on the floor, legs uncrossed. Arm was supported at heart level, slightly bent with palm up. Manometer was positioned at eye level of student who was measuring BP. Patient was instructed not to talk. The brachial artery was palpated and center of the cuff’s bladder was positioned over it. The cuff was evenly and snugly applied one-inch above the bend of the arm. For estimation of systolic blood pressure, the radial arterial pulse was palpated; the cuff was inflated to the point where the pulse can no longer be felt. The cuff was deflated slowly, noting the point where the pulse can be felt (this is the estimated systolic bp) and the cuff was deflated rapidly. The maximum inflation level (MIL) was measured by adding 20-30 mmHg to the estimated systolic pressure (this is the level the cuff should be inflated to when taking the bp measurement). After determining the MIL, and then 15-30 seconds were waited before re-inflating the cuff. For taking the BP measurement, earpieces of the stethoscope are angled forward to fit snugly. The bell or the diaphragm head of the stethoscope was placed lightly over the brachial artery at the bend of the elbow, but with good skin contact as too much pressure can close off the vessel and distort the sounds. Inflate the cuff to the MIL rapidly and steadily. The air in the cuff was released so the pressure falls at 2-3 mmHg per second. The first of two consecutive beats were noted that appeared in relation to the number on the gauge (this is the systolic pressure). Deflation continued and was noted on the gauge where the last sound was heard. This is the diastolic pressure. Deflation continued for 10 mmHg past the last sound as this assures that the absence of sound is not a skipped beat but is the true end of the sound, then the cuff was deflated rapidly and completely. The second measurement was taken after a minute. The average of those readings was used.(8)

For hand washing, any jewellery should be removed first and sleeves are pushed above the wrists. The water tap was turned on and hands were done wet thoroughly, keeping hands and forearms lower than elbows. A palm-sized amount of hand soap was applied. The hand hygiene was performed using plenty of lather and friction for at least 15 seconds, hands are rubbed palm to palm, back of right and left hand (fingers interlaced), palm to palm with fingers interlaced, rotational rubbing of left and right thumbs, fingertips are rubbed against the palm of opposite hand, wrists are rubbed, and sequence was repeated at least 2 times. The fingertips are clean pointing downward throughout. The hands are washed for a minimum of 20 seconds. The hands and forearms are kept lower than elbows during the entire washing. Hands are rinsed with water keeping fingertips pointing down, so water runs off fingertips. Water should not be shaken from hands and do not lean against the sink or touch the inside of the sink during the hand-washing process. After that hands are dried thoroughly from fingers to wrists with a paper towel or air dryer and the paper towel(s) are disposed off. Later a new paper towel is used to turn off the water tap and it is disposed off (9).

Students’ self-confidence (10,11) was measured for both groups using tool three, after performing the procedure.

Effectiveness of training was measured using Likert scale (11).

Students’ perception level (11) was measured for both groups of students after performing the procedure and the total score was calculated using tool four.

For Students’ self-confidence and Students’ perception level tool(10), it consisted of 16 statements with a five-point Likert scale ranged from strongly disagree (1) to strongly agree (5). The higher the score, the higher the satisfaction level. The total score ranged from “16-80”. The cut off point for “High satisfaction” is ≥75% of the total score; “moderate satisfaction” is between 50% to less than 75% of the total score, while “low satisfaction” less than 50% of the total score. It represented as follows; low satisfaction (16-39), moderate satisfaction (40-59), and high satisfaction (60-80).

**Table: Likert scale (11)**

| **Table: Likert scale (11)** |  |  |  |  |
| --- | --- | --- | --- | --- |
| 1 | 2 | 3 | 4 | 5 |
| Never | Rarely | Sometimes | Often | Always |
| Completely disagree | Disagree | Neutral | Agree | Completely agree |

### Study tool/s

The validity of study tools was determined by carrying out pilot study among 10 subjects. Checking of the check list and reliability of the study tool was determined by using cronbach’s alpha coefficient test value degree 0.92.

### Data collection process

A common scoring checklist for outcome evaluation was prepared to evaluate the student’s skills following the training session to avoid discrepancies in scoring among the faculties.

Pre-test and post-test were conducted to enquire about effectiveness of teaching method used and feedback.

## RESULTS

In the control group 52% students were able to measure blood pressure and in study group 88% of students were able to do following skill sessions (table 2).

**Table 1:** Blood pressure measurement (Procedure 1)

| Group |  | Frequency | Percent | P value |
| --- | --- | --- | --- | --- |
| Control Group | Scores <80% | 12 | 48.0 | 0.005* |
|  | Scores ≥80% | 13 | 52.0 |  |
|  | Total | 25 | 100.0 |  |
| Study Group | Scores <80% | 3 | 12.0 |  |
|  | Scores ≥80% | 22 | 88.0 |  |
|  | Total | 25 | 100.0 |  |

**Table 2:**
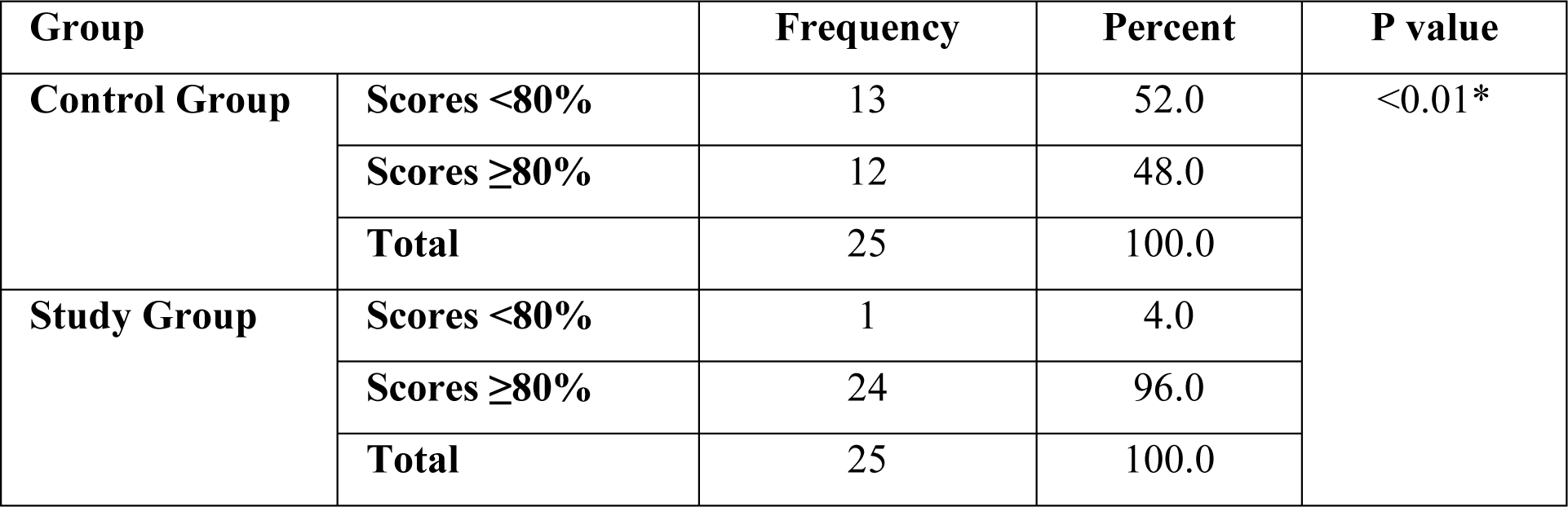
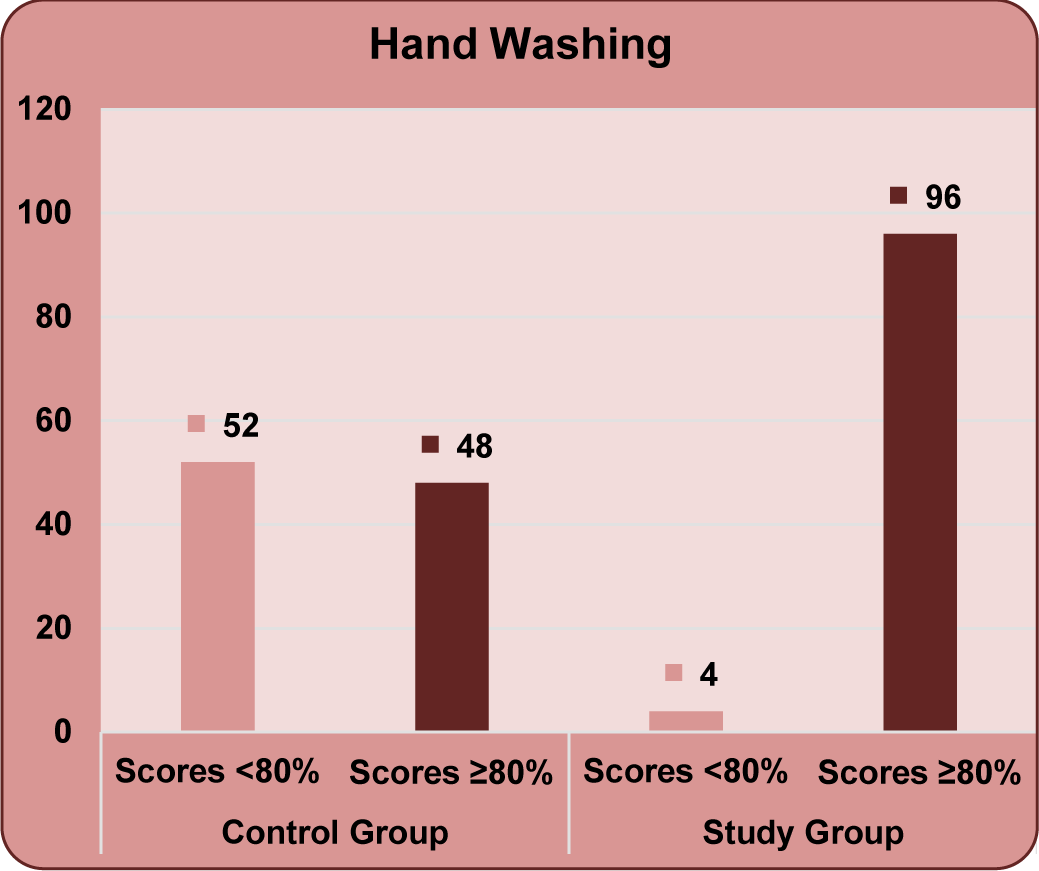
Hand Washing (Procedure 2)

**Table 3:**
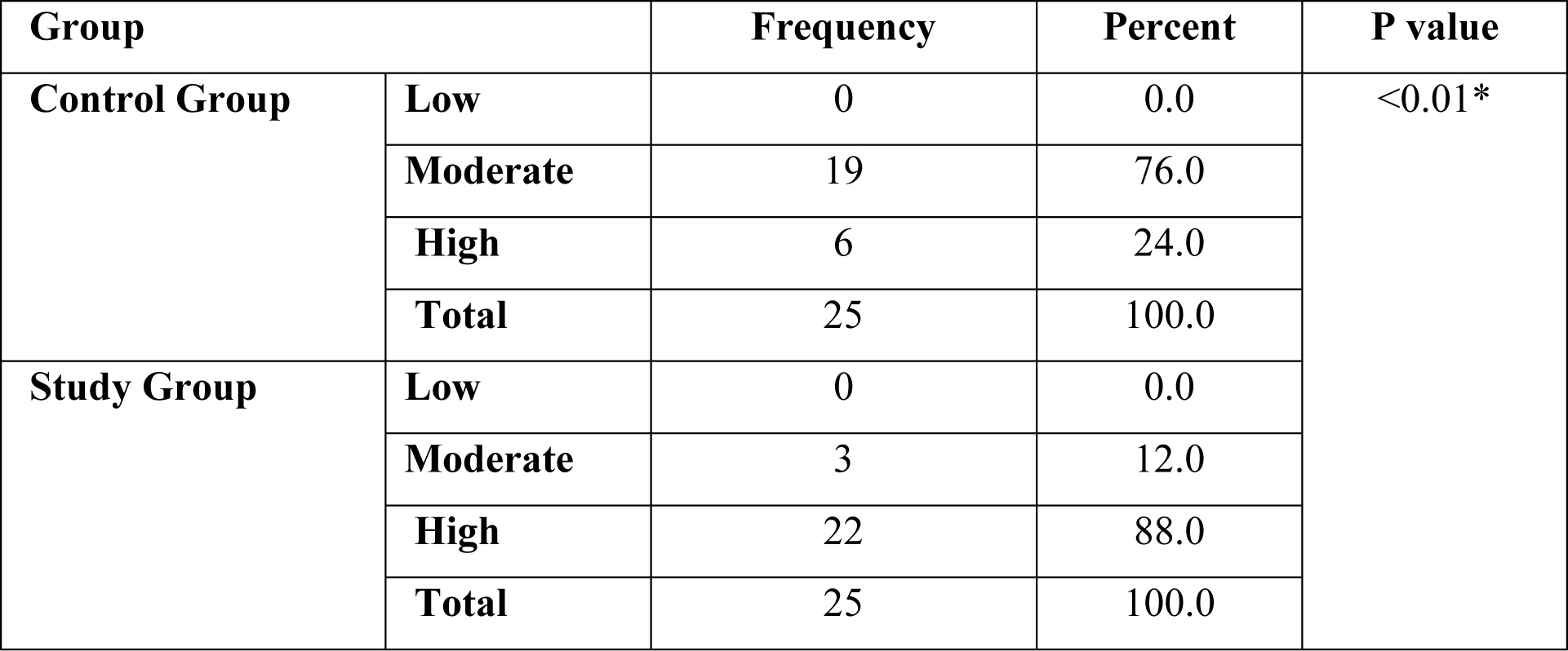
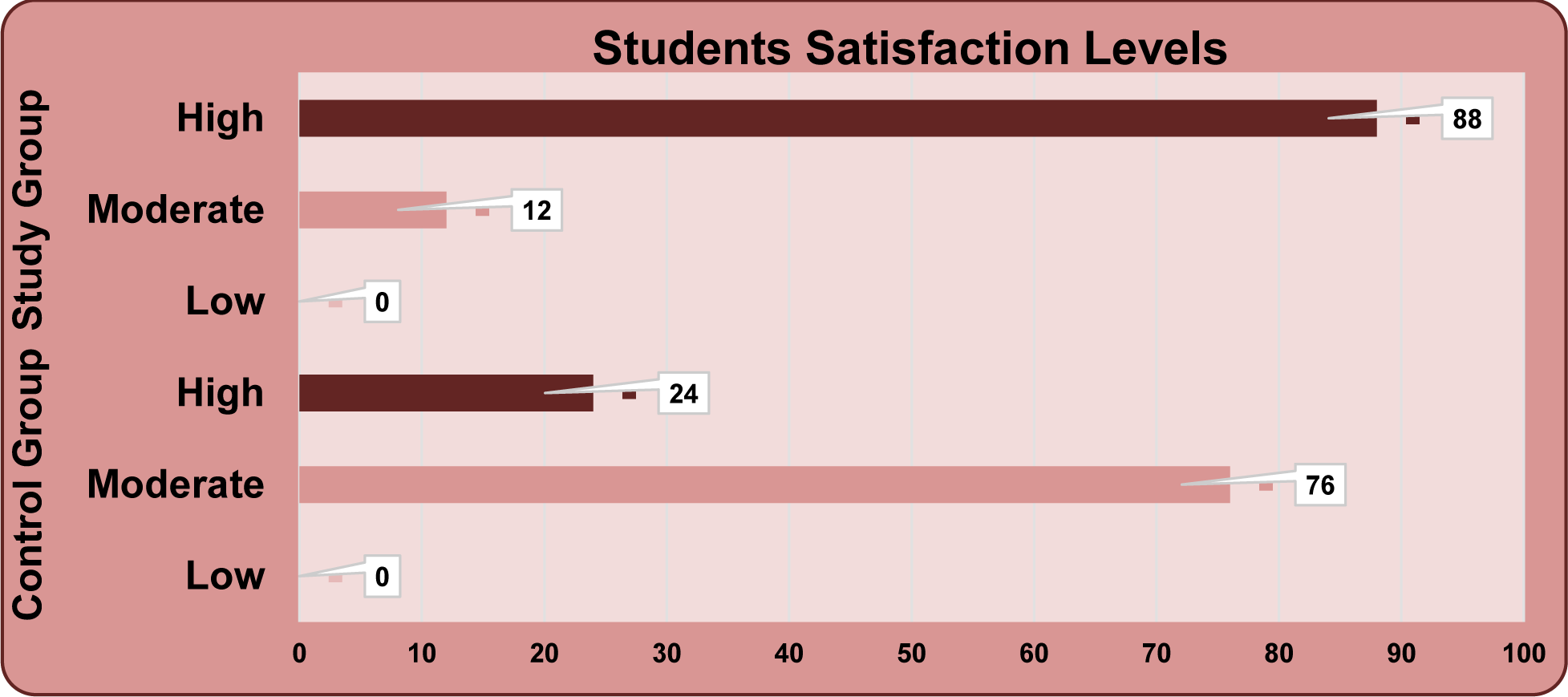
Students self-confidence and satisfaction levels.

Regarding Students self-confidence and satisfaction levels, 24% were highly satisfied and confident in the control group as comparative to 88% students among study group (table 4).

**Table 4:** Effectiveness of training.

|  | Control Group |  | Study Group |  | P value |
| --- | --- | --- | --- | --- | --- |
| Effectiveness of training: Below Average | 3 | 12% | 0 | 0% | <0.01* |
| Effectiveness of training: Average | 10 | 40% | 1 | 4% |  |
| Effectiveness of training: Good | 9 | 36% | 2 | 8% |  |
| Effectiveness of training: Very Good | 2 | 8% | 21 | 84% |  |

Regarding average and very good effectiveness of training, a statistical difference was found between control and study group.

**Table 5:** Frequency distribution as per Student perception of control and study group.

| Student perception | Control Group |  | Study Group |  | P value |
| --- | --- | --- | --- | --- | --- |
| Interest | 2 | 4% | 0 | 0% | <0.01* |
| Interactive | 3 | 12% | 0 | 0% |  |
| Time consuming | 3 | 12% | 0 | 0% |  |
| Understanding | 9 | 36% | 1 | 4% |  |
| Recall | 3 | 12% | 2 | 8% |  |
| Confidence | 4 | 16% | 21 | 84% |  |

## DISCUSSION

Peyton’s approach is a combination of various aspects of the learning theory. It is comprised of four steps including demonstration, deconstruction, comprehension and execution. Step 1 and 2 are based on social-cognitive learning theory while step 4 is based on behaviorists learning theory (12). The present study was undertaken to compare the effectiveness of training effect of peers’ application of modified Peyton’s four-step approach versus traditional learning on medical students’ performance to measure blood pressure and performing hand washing procedures. The students’ self-satisfaction and confidence level in study group and control group was also compared in the current study.

It was found that in the control group 52% students were able to measure blood pressure and in study group 88% of students were able to do following skill sessions and in the control group 48% students were able to wash hands as per required protocol and in study group 96% of students were able to do following skill sessions. Similar to our study, Mohammed AA et al conducted a like study among nursing students by applying modified Peyton’s four-step approach by peers and reported it as an innovative clinical teaching method that has a positive effect on the clinical performance of the pediatric nursing students immediately after teaching as they exhibit higher competency in performing neonatal CPR procedure than students who taught through traditional learning (10). Another similar study by Sabaq AG et al supported in favor of the peer teaching approach evidence by an obvious improvement in students’ knowledge and performance scores and authors reported that majority of students in the study group felt less anxious, comfortable, more self-confident in teaching as well as revealed improvement in their communication skills (13).

A peer is a student of the same age, group, academic level, or experience level. The Oxford Dictionary (2009) defines a “peer” as someone of the same age or someone who was attending the same university. The term “peer” can also refer to people who have equivalent skills of different experiences. Peer learning is also described as a two-way reciprocal learning activity which includes sharing knowledge, ideas, and experiences in a way that has some benefits for both groups of peer and student (7). Regarding students self-confidence and satisfaction levels in the present study, 6% were satisfied and confident in the control group as comparative to 22% students among study groug and a statistical difference was noted in confidence level in between two groups. Regarding average and very good effectiveness of training, a statistical difference was found between control and study group. Similar to our study, Mohammed AA et al reported that modified Peyton’s four-step approach contributed in improvement of the students’ satisfaction level in performing (10). Another corresponding study conducted by Zentz SE et al over a period 2 years in which 342 students participated in peer-assisted learning, major outcomes identified by sophomores were reduced anxiety and increased confidence. A major benefit for seniors was reflection on their professional development, which strengthened their confidence and facilitated transition into the role of professional nurse and this peer-assisted learning as an effective teaching strategy for learning skills and implementing the roles of the medical professional were supported by their study (14). Different roles for medical students can be developed within a peer-assisted learning scheme, presenting different challenges and benefits to participants (15).

The **limitations** of present study are that only two activities were used for teaching and assessment of SPSS skill learning. More activities should be used to gather more evidence. Also, reassessment of acquired skills should be at multiple time intervals in order to assess the long-term retention of skills.

## CONCLUSIONS

Students who learnt through their peers using modified Peyton’s four–step approach showed rapid learning, more confidence and exhibited higher competency in performing blood measurement procedure and hand washing techniques than those who learned through traditional method.

## Data Availability

All data produced in the present study are available upon reasonable request to the authors

## DISCLAIMER

This research project was supported in part by the Foundation for Advancement of International Medical Education and Research. However, the findings and conclusions do not necessarily reflect the opinions of this organization.

## ACKNOWLEDGEMENTS

The study was developed and implemented as a part of CMCL-FAIMER fellowship for the first author. Hence the team thanks faculty and fellows of Christian Medical College, Ludhiana FAIMER Regional institute who contributed immensely to improving its methodological quality by providing continuous feedback and guidance.

## REFERENCES

1. Halsted WS. The training of the surgeon. Bull Johns Hop Hosp. 1904:267–75.

2. Giacomino K, Caliesch R, Sattelmayer KM. 2020. The effectiveness of the Peyton’s 4-step teaching approach on skill acquisition of procedures in health professions education: A systematic review and meta-analysis with integrated meta-regression. PeerJ 8:e10129 https://doi.org/10.7717/peerj.10129

3. Herrmann-Werner A, Nikendei C, Keifenheim K, Bosse HM, Lund F, Wagner R, Celebi N, Zipfel S, Weyrich P. “Best practice” skills lab training vs. a “see one, do one” approach in undergraduate medical education: an RCT on students’ long-term ability to perform procedural clinical skills. PLoS One. 2013;8(9):e76354.

4. Krautter M, Weyrich P, Schultz JH, Buss SJ, Maatouk I, Junger J, Nikendei C. Effects of Peyton’s four-step approach on objective performance measures in technical skills training: a controlled trial. Teach Learn Med. 2011;23(3):244–50.

5. Peyton JWR. Teaching in the theatre. In: Peyton JWR, editor. Teaching and learning in medical practice. Manticore Europe; 1998: p. 171–80.

6. Ahmed FR, Morsi SR, Mostafa HM. Effect of Peyton’s Four Step Approach on skill acquisition, self-confidence and self-satisfaction among critical care nursing students. IOSR-JNHS. 2018;6:38–47.

7. Ravanipour M, Bahreini M, Ravanipour M. Exploring nursing students’ experience of peer learning in clinical practice. J Educ Health Promot. 2015;4:46. Published 2015 May 19. doi:10.4103/2277-9531.157233

8. Montana Hypertension Initiative. Blood Pressure Measurement Technique Checklist. Last assessed as on 17 July 2022. Available at: https://dphhs.mt.gov/assets/publichealth/Cardiovascular/BPTechniqueChecklistTackleBox.pdf

9. Checklist For hand washing technique: Last assessed as on 17 July 2022. Available at: https://med.libretexts.org/Bookshelves/Nursing/Nursing_Skills_(OpenRN)/04%3A_Aseptic_Technique/4.05%3A_Checklist_for_Hand_Hygiene_with_Soap_and_Water

10. Mohammed AA, Mohammed NY, Ouda MM, Abu ElEla LA. Effect of peers’ application of modified Peyton’s four-step approach versus traditional learning on pediatric Nursing students’ performance. Journal of Health, Medicine and Nursing. 2019 Dec 31;69:65–

11. Sullivan GM, Artino Jr AR. Analyzing and interpreting data from Likert-type scales. Journal of graduate medical education. 2013 Dec;5(4):541–2.

12. Naseem S, Fatima D, Ghazanfar H, Awan SA, Fatima DS. Four step approach by Peyton’s as a tool for teaching SPSS among undergraduate medical students: An experience from Pakistan.

13. Sabaq AG, Farouk M, Ismail SS. Effect of Peer Teaching Versus Traditional Teaching Method on Nursing Students’ Performance Regarding Pediatric Cardiopulmonary Resuscitation. Tanta Scientific Nursing Journal. 2016 May 1;10(1):7–25.

14. Zentz SE, Kurtz CP, Alverson EM. Undergraduate peer-assisted learning in the clinical setting. Journal of Nursing Education. 2014 Mar 1;53(3):S4–10.

15. Hill E, Liuzzi F, Giles J. Peer-assisted learning from three perspectives: student, tutor and co-ordinator. The clinical teacher. 2010 Dec;7(4):244–6.

